# Magnitude, demographics and dynamics of the impact of the first phase of the Covid-19 pandemic on all-cause mortality in 17 industrialised countries

**DOI:** 10.1101/2020.07.26.20161570

**Authors:** Vasilis Kontis, James E Bennett, Theo Rashid, Robbie M Parks, Jonathan Pearson-Stuttard, Michel Guillot, Perviz Asaria, Bin Zhou, Marco Battaglini, Gianni Corsetti, Martin McKee, Mariachiara Di Cesare, Colin D Mathers, Majid Ezzati

**Affiliations:** MRC Centre for Environment and Health, School of Public Health, Imperial College London, London, UK; The Earth Institute, Columbia University, New York, NY, USA; Department of Environmental Health Sciences, Mailman School of Public Health, Columbia University, New York, NY, USA; Population Studies Center, Department of Sociology, University of Pennsylvania, Philadelphia, PA, USA; French Institute for Demographic Studies (INED), Paris, France; Directorate for Social Statistics and Population Census, Italian National Institute of Statistics (Istat), Rome, Italy; Department of Health Services Research and Policy, London School of Hygiene & Tropical Medicine, London, UK; Department of Natural Sciences, Middlesex University London, London, UK; Independent Researcher, Geneva, Switzerland; Regional Institute for Population Studies, University of Ghana, Legon, Ghana

## Abstract

The Covid-19 pandemic affects mortality directly through infection as well as through changes in the social, environmental and healthcare determinants of health^1^. The impacts on mortality are likely to vary across countries in magnitude, timing, and age and sex composition. Here, we applied an ensemble of 16 Bayesian probabilistic models to vital statistics data, by age group and sex, to consistently and comparably estimate the impacts of the first phase of the pandemic on all-cause mortality for 17 industrialised countries. The models accounted for factors that affect death rates including seasonality, temperature, and public holidays, as well as for medium-long-term secular trends and the dependency of death rates in each week on those in preceding week(s). From mid-February through the end of May 2020, an estimated 202,900 (95% credible interval 179,400-224,900) more people died in these 17 countries than would have had the pandemic not taken place. Nearly three quarters of these excess deaths occurred in England and Wales, Italy and Spain, where less than half of the total population of these countries live. When all-cause mortality is considered, the total number of deaths, deaths per 100,000 people, and relative increase in deaths were similar between men and women in most countries. Further, in many countries, the balance of excess deaths changed from male-dominated early in the pandemic to being equal or female-dominated later on.

Taken over the entire first phase of the pandemic, there was no detectable rise in all-cause mortality in New Zealand, Bulgaria, Hungary, Norway, Denmark and Finland and for women in Austria and Switzerland (posterior probability of an increase in deaths <90%). Women in Portugal and men in Austria experienced relatively small increases in all-cause mortality, with posterior probabilities of 90-99%. For men in Switzerland and Portugal, and both sexes in the Netherlands, France, Sweden, Belgium, Italy, Scotland, Spain and England and Wales, all-cause mortality increased as a result of the pandemic with a posterior probability >99%. After accounting for population size, England and Wales and Spain experienced the highest death toll, nearly 100 deaths per 100,000 people; they also had the largest relative (percent) increase in deaths (37% (95% credible interval 30-44) in England and Wales; 38% (31-44) in Spain). New Zealand, Bulgaria, Hungary, Norway, Denmark and Finland experienced changes in deaths that ranged from possible slight declines to increases of no more than 5%. The large impact in England and Wales stems partly from having experienced (together with Spain) the highest weekly increases in deaths, more than doubling in some weeks, and having had (together with Sweden) the longest duration when deaths exceeded levels that would be expected in the absence of the pandemic.

The heterogeneous magnitude and character of the excess deaths due to the Covid-19 pandemic reflect differences in how well countries have managed the pandemic (e.g., timing, extent and adherence to lockdowns and other social distancing measures; effectiveness of test, trace and isolate mechanisms), and the resilience and preparedness of the health and social care system (e.g., effective facility and community care pathways; minimising spread of infection within hospitals and care homes, and between them and the community).

---

The Covid-19 pandemic has led to hundreds of thousands of deaths directly among infected patients. The pandemic has also profoundly changed the social, economic, environmental and healthcare determinants of morbidity and mortality. These changes impact health through a number of routes^1^ including denied or delayed disease prevention and medical procedures for acute and chronic care; loss of job and income; disruption of social networks; changes in crime and self-harm; changes in quantity and quality of food, and the use of tobacco, alcohol and other drugs; and changes in road traffic crashes, other injuries and air pollution as a result of reduced and modified mobility and transportation. How changes in these social, environmental and healthcare determinants impact mortality varies across countries based on the extent and timing of the epidemic and the response, the baseline health of the population, the resilience and agility of the health and social care system, and social and economic safety nets. It is also likely to vary, in both magnitude and timing, by sociodemographic characteristics. An understanding of the total mortality impacts is needed to understand the true public health impacts of the pandemic and the policy response. Comparative multi-country analyses^2^ help understand how responses can be made more effective and timely, and how health and social care systems may be made more resilient. However, some politicians are rejecting country benchmarking based on the argument that the data, methodology and timing of the analysis are not comparable across countries.^3^ Here, we developed and applied a probabilistic methodology for consistent and comparable quantification of the weekly mortality impacts of the first phase of the Covid-19 pandemic in 16 industrialised countries in central and western Europe plus New Zealand.

## Magnitude of excess deaths

Deaths in all these countries were at the levels that would be expected in the absence of the pandemic through the month of February but started to diverge to higher levels at various times in March in some countries (Fig. 1). From mid-February through the end of May 2020, an estimated 202,900 (95% credible interval 179,400-224,900) more people died in these 17 countries than would have been expected had the pandemic not taken place. This number is larger than the number of deaths from lung cancer in these countries in an entire year, and 2.5 times the number of deaths from diabetes or breast cancer in an entire year.^4^ 103,400 (90,200-114,800) of these deaths were in men and 99,600 (83,900-115,000) in women (Extended Data Table 1). In relative terms, this amounts to an 18% (16-20) increase in deaths over this period in these countries together. Italy, Spain and England and Wales accounted for 23%, 23% and 28% of these excess deaths, respectively.

**Fig. 1.**
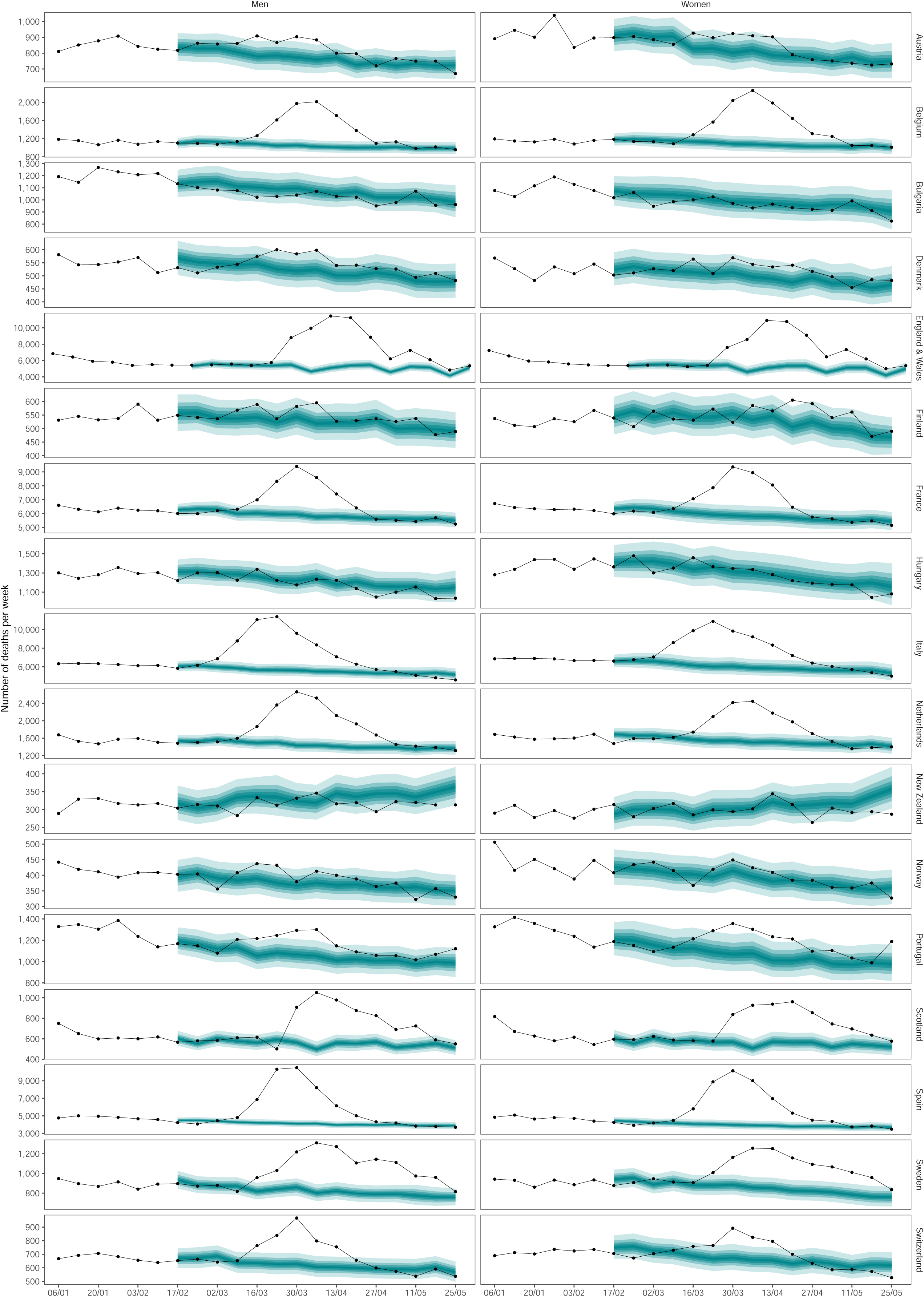
Weekly number of deaths from any cause from January 2020 through May 2020. The points show reported deaths (placed at the start of each week in this graph). The turquoise-shaded areas show the predictions of how many deaths would have been expected from mid-February had Covid-19 pandemic not taken place. The shading shows the credible intervals around the median prediction, from 5% (dark) to 95% (light) in 10% increments. See Extended Data Fig. 1 for results by age group.

Posterior probability measures the extent to which an estimated change in deaths is likely to be a true increase or decrease. It would be 50% in a country and/or week in which an increase is statistically indistinguishable from a decrease, and larger posterior probability indicates more certainty in an increase. The posterior probability that there was a rise in deaths over the entire first phase of the pandemic was <50% (i.e., a decline in deaths is more likely than an increase) for both sexes in New Zealand, Bulgaria and Hungary; 50-75% for women in Norway and Austria; 75-90% for men and women in Denmark and Finland, men in Norway and women in Switzerland; 90-99% for women in Portugal, and men in Austria; and >99% for men in Portugal and Switzerland and for both sexes in the Netherlands, France, Sweden, Belgium, Italy, Scotland, Spain and England and Wales (Fig. 2). In countries and sexes where mortality increased relative to the no-pandemic baseline with a posterior probability of at least 90%, the number of excess deaths per 100,000 people was lowest in Austria (15.0, −0.2-29.6), Switzerland (21.5, 6.7-34.4) and Portugal (27.7, 4.0-50.1) for men, and in Portugal for women (28.0, 0.7-54.4) (Fig. 2). It was highest in Spain and England and Wales, with posterior median estimates for the two sexes ranging from 90 to 102 per 100,000 population. The posterior median increase was also >50 per 100,000 people in Sweden, Belgium, Italy and Scotland. Relative increase in deaths, compared to what would be expected in the absence of the pandemic, ranged from <10% in Austrian, Swiss and Portuguese men and Portuguese women to one quarter or more in Belgium, Italy, Scotland, Spain and England and Wales (Fig. 3). The largest rise in mortality for men was most likely to be in England and Wales (66% posterior probability of having the largest percent increase, and 59% of having the largest number of deaths per 100,000 people) followed by Spain; for women, Spain was most likely to have experienced the largest rise in mortality (62% posterior probability of having the largest percent increase and 55% of having the largest number of deaths per 100,000 people) followed by England and Wales.

**Fig. 2.**
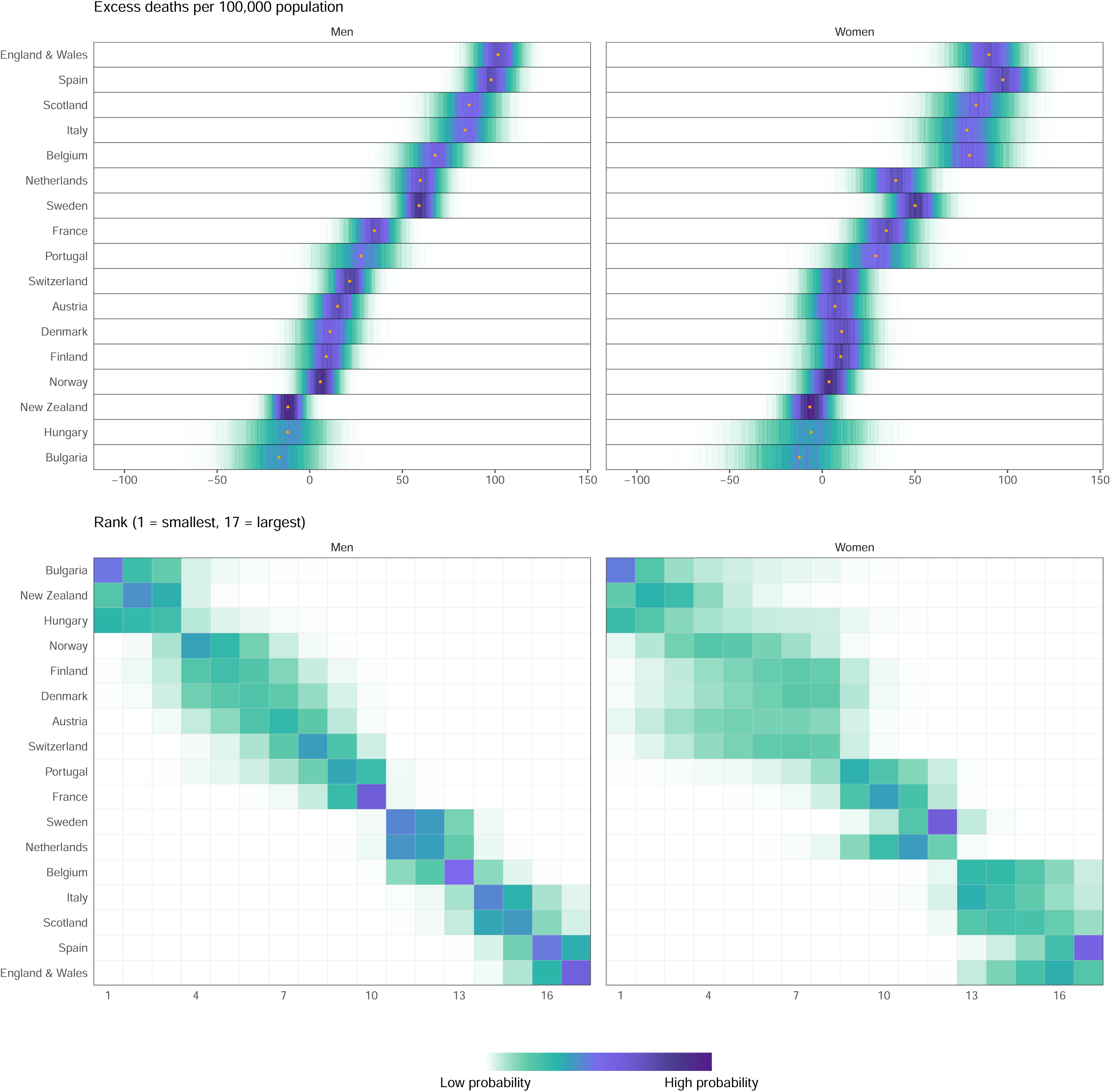
(A) Posterior distribution of excess deaths from any cause per 100,000 people from mid-February to end of May 2020. Gold dots show the posterior medians. (B) Probability distribution for the country’s rank. Countries are ordered vertically by median increase from smallest (at the bottom) to the largest (at the top) in men. See Extended Data Fig. 2 for results by age group.

**Fig. 3.**
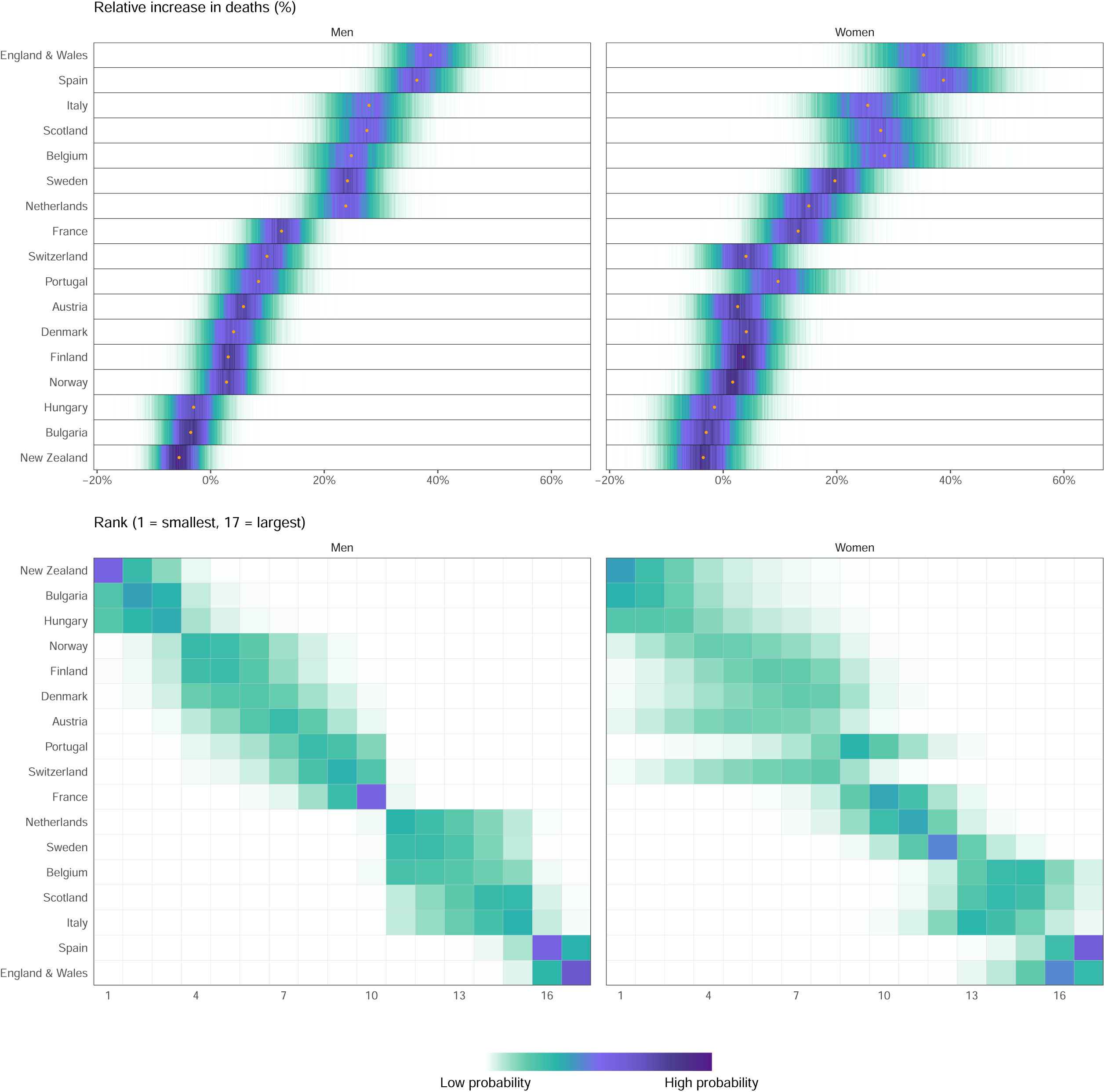
(A) Posterior distribution of percent increase in deaths from any cause from mid-February to end of May 2020. Gold dots show the posterior medians. (B) Probability distribution for the country’s rank. Countries are ordered vertically by median increase from smallest (at the bottom) to the largest (at the top) in men. See Extended Data Fig. 3 for results by age group.

Taken across all countries, the number of excess deaths from all causes was 23% higher than the number of deaths assigned to Covid-19 as underlying cause of death (Extended Data Table 1). The difference between all-cause excess and Covid-19 deaths was largest in Spain and Italy, where all-cause excess deaths were 68% and 39%, respectively, higher than deaths assigned to Covid-19. This difference is likely due to a combination of undetected infections,^5,6^, whether deaths from “suspected infections” (i.e., based on clinical symptoms) are assigned to Covid-19 or not^7^, and some increase in mortality from other diseases due to reductions in acute and chronic care^8-14^. In contrast to Italy and Spain, the overall (all-cause) number of excess deaths was smaller than deaths assigned to Covid-19 in France and Belgium^7,15,16^. As a result, while France and Spain have reported similar number of Covid-19 deaths, all-cause mortality increased by twice as much in Spain as in France. These variations show the importance of measuring the death toll due to the pandemic using all-cause mortality.

## Timing of excess deaths

Italian men were the first group to experience a rise in mortality, in the 1^st^ week of March 2020 with a posterior probability of >90%, followed by Italian women and Spanish men in the subsequent week (Fig. 4). Deaths in some countries with large early mortality, e.g., Italy and France, returned to levels that would be expected in the absence of the pandemic in April, followed by those in Spain. Deaths remained above the levels expected in the absence of the pandemic in England and Wales and Sweden for most/all of May which resulted in longer periods of adverse impact. As a result, in countries and sexes where the posterior probability of an increase in deaths was >90%, the period of time when deaths were higher than would be expected in the absence of the pandemic ranged from five weeks in Austrian men to 9-11 weeks in men and women in England and Wales and Sweden.

**Fig. 4.**
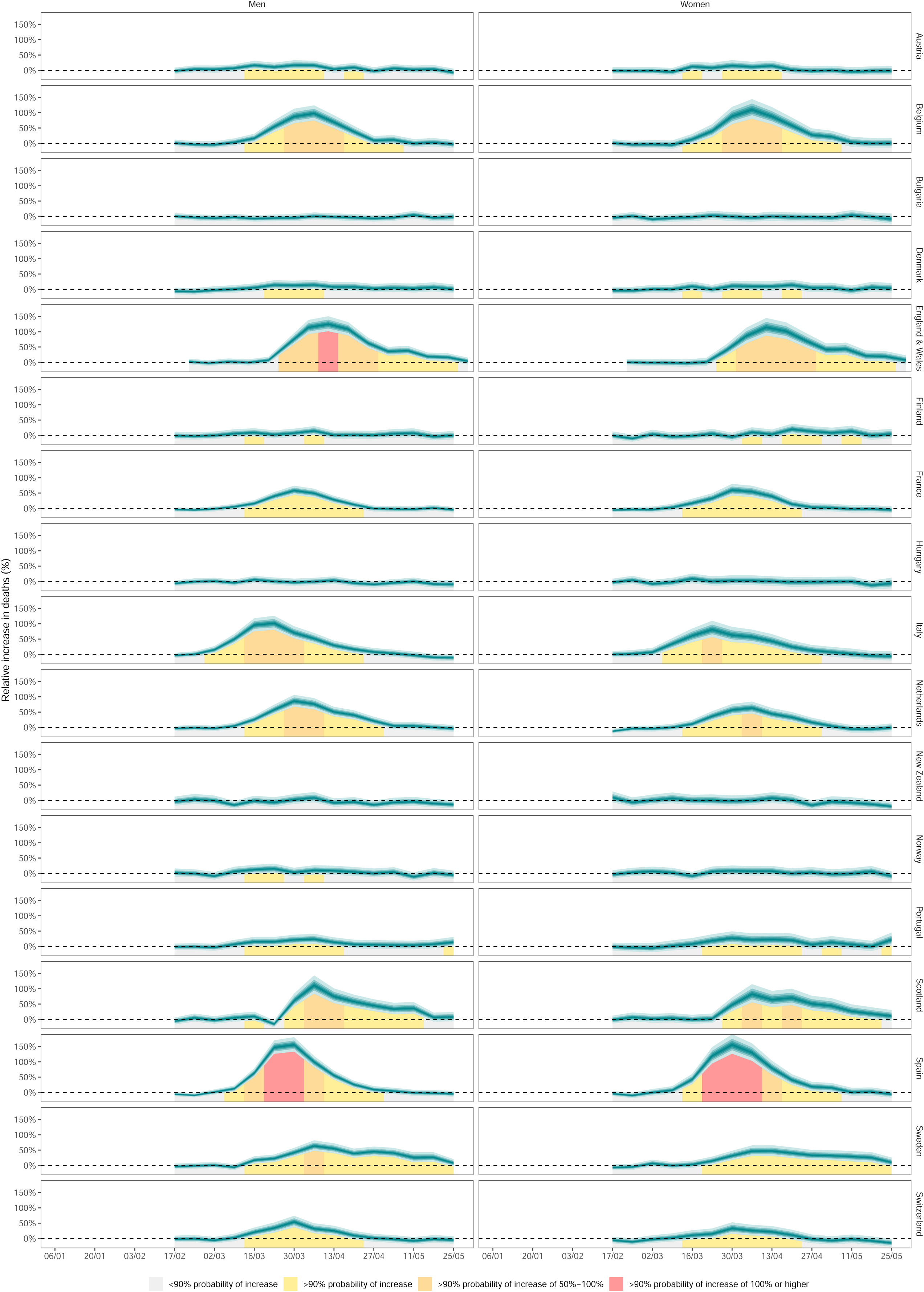
Weekly percent increase in mortality from any cause as a result of the Covid-19 pandemic by country. The turquoise shading shows the credible intervals around the median prediction, from 5% (dark) to 95% (light) in 10% increments. The background shading indicates the magnitude of the weekly increase that was detectable with a posterior probability of at least 90%. See Extended Data Fig. 4 for results by age group.

The large adverse impact of the pandemic in England and Wales and to some extent in Spain is a consequence of both having long durations and large weekly rises, with a > 90% posterior probability that for some weeks deaths in men and women in Spain and men in England and Wales more than doubled. In contrast, Portugal, Switzerland, and possibly France had smaller weekly rises, and for fewer weeks, and hence had overall increases between one quarter and one half of those in England and Wales and Spain. Sweden had the longest duration of excess deaths, but had smaller weekly increases in deaths than countries like England and Wales, Spain, Scotland, Italy and Belgium. As a result, the overall mortality toll in Sweden, in terms of relative increase and deaths per 100,000 people, fell between those of low-moderate-impact countries (e.g., Portugal, Switzerland) and places with extreme tolls (e.g., Spain and England and Wales).

## Demographic distribution of excess deaths

Although it is widely quoted that more men die from Covid-19 infection,^17,18^ the number of excess deaths for all causes, excess deaths per 100,000 people, and relative increase in deaths were similar between men and women in most countries (Fig. 4). In all 17 countries together, 103,400 (90,200-114,800) men died from any cause of death as a result of the pandemic compared to 99,600 (83,900-115,000) women. Further, in many countries the balance of excess deaths changed from male-dominated early in the pandemic to being equal (e.g., England and Wales) or female-dominated (e.g., Italy, Spain and France) later on.

When considered in terms of relative increase in deaths, male disadvantage was largest in the Netherlands (24% (16-32) increase in male deaths compared to 15% (7-24) in female deaths) and Switzerland (10% (3-17) increase in males; 4% (-4-12) in females). In contrast, in Belgium (25% (16-34) increase in males; 28% (18-40) in females) and Spain (36% (29-44) increase in males; 39% (29-50) in females) there was a slight female disadvantage in total mortality impacts. A male disadvantage in pandemic-related excess deaths was more pronounced before 65 years of age, whereas in older ages the relative impacts were similar between men and women (Fig. 5). For example, the pandemic led to an estimated 18% (9-28) increase in deaths in males younger than 65 years compared to 2% (-7-11) in females of the same age in Sweden; 14% (7-20) and 7% (1-14) in Italy; and 11% (5-17) and -2% (-8-4) in the Netherlands.

**Fig. 5.**
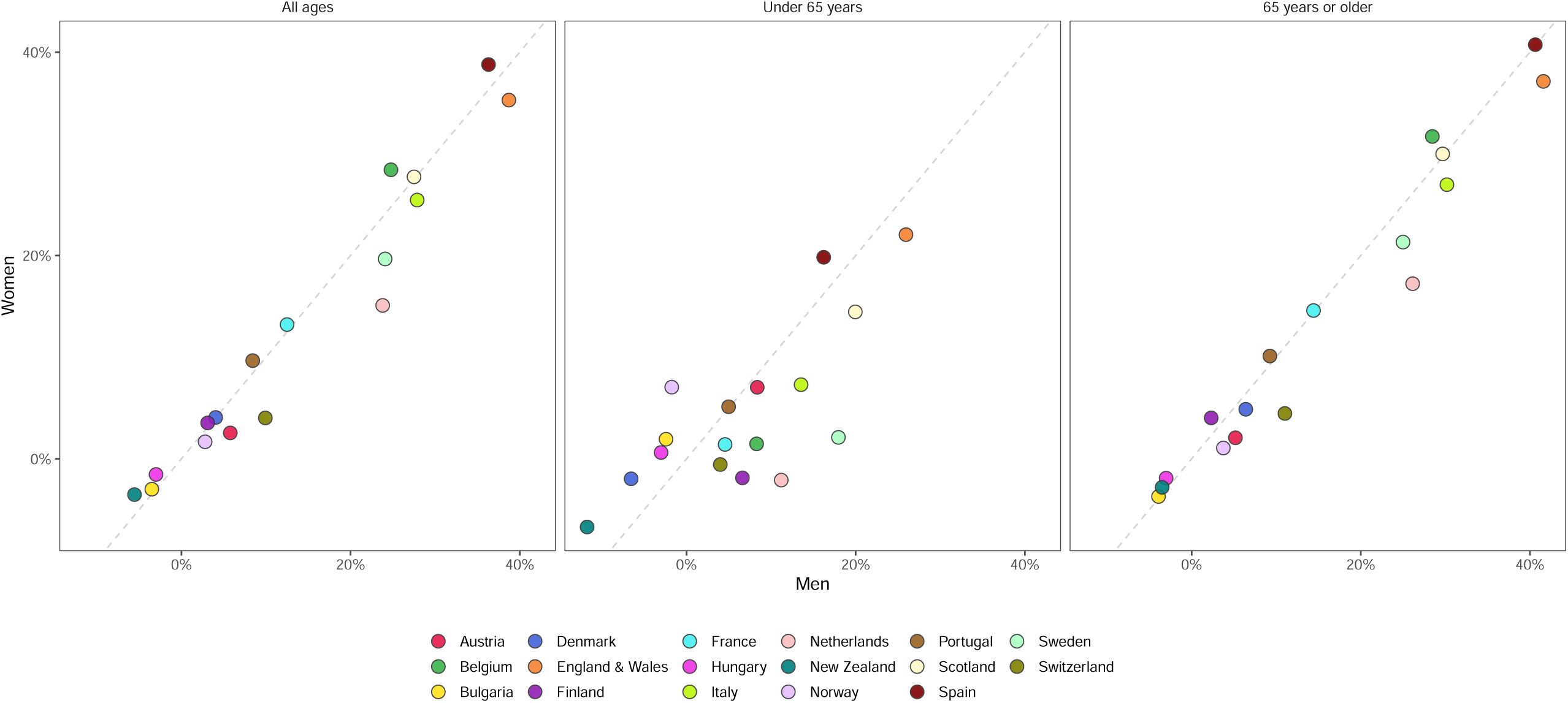
Comparison of percent increase in deaths from any cause as a result of the Covid-19 pandemic between men and women, for all ages and by age group.

In absolute terms, the total mortality toll of the pandemic was overwhelmingly in those aged 65 years and older, who experienced 93% of all excess deaths due to the pandemic. In relative terms, older people were also impacted more, with mortality in these ages being >40% higher than it would have been in the absence of the pandemic in Spain and England and Wales, and ∼30% in Belgium, Scotland and Italy. The largest impact on those younger than 65 years was in England and Wales, 27% (20-34) for males and 23% (17-29) for females, followed by Scotland, Spain, Sweden and Italy. In men and women in New Zealand, and men in Denmark, there may have been a slight decline in deaths in men aged <65 years as a result of the pandemic, with posterior probabilities of the observed declines being true declines >90%. In these ages, injuries are an important cause of death especially for men. For example, men younger than 65 years in New Zealand, the only group with a 99% posterior probability of a decline in total mortality, have a higher share of all deaths that are from injuries than in other countries in this analysis^4^.

## Lessons from heterogeneous mortality impacts

With our consistent and comparable analysis, we have identified four groups of countries in terms of the overall death toll of the first phase of the Covid-19 pandemic: the first are those that have avoided a detectable rise in all-cause mortality, and include New Zealand, Bulgaria, Hungary and Norway. The second and third groups of countries experienced a low-medium impact of the pandemic on overall deaths and include Denmark, Finland, Austria, Switzerland and Portugal (low impact) and France, the Netherlands and Sweden (medium impact). The fourth group, which experienced the highest mortality toll, consists of Belgium, Italy, Scotland, Spain and England and Wales.

The total death toll for the first phase of the Covid-19 pandemic in a country is affected by three key groups of determinants: the characteristics of the population and communities they live in, the response policies that affect mortality positively by interrupting transmission and negatively by isolation and denial of essential services, and upstream health systems and social and political factors that shape these policies^19^. The first group of determinants are contextual characteristics of individuals and communities that make them vulnerable or resilient to the spread of infection and the adverse consequences of restrictions, and include baseline demography and health, social networks and inequalities, employment and occupation, and environmental features such as transport and housing. The risk of death from Covid-19 increases with age, social and material deprivation, and in the presence of long-term conditions such as obesity, hypertension, diabetes and vascular and kidney diseases. Most countries in our analysis have an aging population and none stands out as particularly older or younger than others. For example, the share of population who are 65+ years ranges from 16% in New Zealand to 23% in Italy and Portugal, but this share was barely correlated with excess mortality (correlation coefficients <0.15). Further, our age-specific results are not affected by age differences across countries. Obesity and diabetes are higher and hypertension is lower in the UK, which experienced one of the highest impacts, than in other European countries in our analysis.^20-22^ But New Zealand, which had no detectable excess deaths, has an even higher prevalence of obesity and diabetes than the UK whereas Belgium, Italy and Spain, which have lower prevalence, also experienced large impacts. Similarly, although reported multi-morbidity varies across Europe,^23^ these are not correlated with excess mortality: Sweden and Denmark, which had vastly different excess deaths, have low levels of multi-morbidity; Hungary, Spain and Italy, which also span the entire range of excess mortality, have some of the highest. Finally, although the UK has higher relative poverty than countries like Norway, Denmark and Finland,^24^ excess deaths were higher in Sweden (similar relative poverty to Denmark and Finland) than in New Zealand (similar relative poverty to the UK). In summary, although these factors are undoubtedly important in terms of the magnitude of the mortality impact, they alone are unlikely to account for the massive cross-country variation observed here. Other important population characteristics remain unexplored and require consistent data. For example, in some countries, regional outbreaks have started among low-wage workers in poor working conditions, such as garment factories and food processing plants. The role of overcrowded social housing complexes and public transportation (and more generally extent and means of mobility)^25^ in the extent and geographical distribution of transmission is also unknown.^26^

The second determinant of mortality toll of the pandemic is the policy and public health response which has varied vastly across countries in timing, character and extent.^27^ The timing of lockdown in relation to when initial infections occurred^28^ affects the peak number of people who are infected, which drives both the number of deaths from Covid-19 and the pressure on the healthcare system that displaces routine care for other diseases. The stringency of the lockdown, together with the extent and effectiveness of testing, contract tracing and isolation, determines how long it takes for the number of cases to return to low absolute levels, and can therefore account for some of the variations in the intensity and duration of excess deaths observed here. Among the countries analysed here, New Zealand, Hungary, Norway and Finland acted early in terms of putting in place various movement restrictions or lockdown^27,29^ and kept the number of cases to such low levels that they could identify and isolate cases and their contacts through their existing public health system. Austria and Denmark experienced an early rise in the number of cases but enacted lockdowns soon after and used widespread and effective testing, tracing and isolation to contain the epidemic and its mortality impact. At the other extreme, Italy, which was the initial European epicentre of the pandemic, Spain, the Netherlands, France and the UK, put lockdown measures in place only after the number of cases and deaths had risen to levels that the epidemic continued for weeks. For example, the UK, Spain, Italy, France and the Netherlands introduced a lockdown after a larger number of cases had been detected, and after a longer period since the first few Covid-19 deaths occurred, than New Zealand and other countries in Europe such as Denmark^27,28,30^. Once in place, the lockdown was more stringent in Spain, Italy and France than in the UK and Belgium.^30^ Countries also varied in how extensively they conducted community testing, contact tracing and isolation of cases and their contacts, with Austria, Denmark, Finland, New Zealand and Norway introducing extensive and effective systems and Belgium, Spain, France and the UK being more limited in community testing and/or contact tracing^27,31^.

Third, the preparedness and resilience of the public health infrastructure not only influence how well the spread of infection is controlled, but also could feed back into the choice of policy. Denmark and Austria (as well as Germany for which data were not available for our analysis) were able to scale up testing rapidly because they had extensive laboratory networks and public health infrastructure in place. Some central European countries had existing contact tracing infrastructure, a legacy of their more recent experience with infectious diseases such as tuberculosis. Others had more limited capacity, such as New Zealand’s contract tracing system, but were able to scale it up rapidly based on the existing public health structures. In contrast, the UK and Spain had less testing capacity (or ability to utilise capacity in non-governmental labs early in the epidemic) and weaker contact tracing and isolation systems, especially early in the pandemic, with largely fragmented strategies and information flow^32^. Countries also vary substantially in terms of how their health care system continued to provide life-saving services: those countries that had less capacity and were less able to rapidly enhance capacity, partly related to uneven health and social care spending, responded less effectively to health care needs. Notably, per capita spending is lower in the UK, Italy and Spain than in Austria, Norway, Sweden and Denmark^33^. A visible impact of financing variation is on the number of hospital beds, which on a per-capita basis in Austria is nearly three times that of the UK^34^. Beyond providing life-saving services, the spread of infection within hospitals and care homes, and between them and community, is itself an important determinant of infection in both the general population and vulnerable groups^35,36^. Where hospital beds (and more specifically intensive care beds) are more limited, e.g., in the UK and Spain, concerns about breeching capacity may have led to delaying admission of Covid-19 and other patients until their health deteriorated, and to early discharge of patients to care homes without systematic testing. Care homes also contributed to large numbers of deaths in Belgium, France and Sweden^35^. Less visible is the variation in community-based and primary care which affect preventive and pre-hospital care for Covid-19 patients as well as for other conditions that either increase vulnerability to poor outcomes from the virus or can themselves lead to death.

The simplest lesson of these vast differences in excess deaths associated with the first phase of the pandemic is that, should subsequent waves of the pandemic occur, immediate lockdowns may be needed^37^ to avoid large number of excess deaths seen in countries such as England and Wales, Italy and Spain. Timely lockdowns in turn require effective surveillance and agile operational systems, with sufficient geographical granularity to limit restrictions to as small an area as possible. Lockdowns, while effective for suppressing transmission, have substantial adverse short- and long-term health, psychosocial and economic impacts^38-40^. Lockdowns can be avoided altogether or be a mechanism of last resort if countries can put in place comprehensive and effective test and trace systems, while engendering a sense of trust and responsibility that increase participation in testing, contact tracing and adherence to isolation advice. Finally, every country needs integrated care pathways at the community and facility level that manage both milder Covid-19 infections and allow other acute and chronic conditions to be rapidly and appropriately triaged and cared for. If we do so, all countries can replicate the low levels of excess deaths in subsequent waves of the pandemic.

## Methods

### Data sources

We included industrialised countries from Europe and the Pacific in the analysis if

- Their total population in 2020 was > 4 million.
- They had up-to-date weekly data on all-cause mortality divided by age group and sex that extended through May 2020. We selected end of May 2020 to have a consistent period of analysis all countries and because our results showed that by this date, the probability that deaths were above the level that would be expected had the pandemic not occurred was within the 90% credible interval in the great majority of countries. We could not obtain age- and sex-specific data for 2020 for Australia, Germany, Greece, the USA, Japan, South Korea and Taiwan, nor for Northern Ireland, and hence they were not included in the analysis.
- The time series of data went back at least to 2015 so that model parameters could be reliably estimated. For countries with longer time series, we used data starting in 2010.

The sources of population and mortality data are provided in Extended Data Table 2. We calculated weekly population through interpolation of yearly population, consistent with approach taken by national statistical offices for intra-annual population calculation^41^. We obtained data on temperature from ERA5^42^, which uses data from global in situ and satellite measurements to generate a worldwide meteorological dataset, with full space and time coverage over our analysis period. We used gridded temperature estimates measured four times daily at a resolution of 30 km to generate weekly temperatures for each first-level administrative region, and gridded population data (https://sedac.ciesin.columbia.edu/data/collection/gpw-v4) to generate population estimates by first-level administrative region in each country. We weighted weekly temperature by population of each first-level administrative region to create national level weekly temperature summaries.

### Statistical methods

The total mortality impact of the Covid-19 pandemic is the difference between the observed number of deaths from all causes of death and the number of deaths had the pandemic not occurred, which is not directly measurable. The most common approach to calculating the number of deaths had the pandemic not occurred has been to use the average number of deaths over previous years, e.g., the most recent five years, for the corresponding week or month when the comparison is made^43^. This approach however does not take into account changes in population size and age structure, nor long- and short-term trends in mortality, which are particularly pronounced for some age groups^44,45^. Nor does this approach account for time-varying factors like temperature, that are largely external to the pandemic, but also affect death rates.

We developed an ensemble of 16 Bayesian mortality projection models that each make an estimate of weekly death rates that would have been expected if the Covid-19 pandemic had not occurred. We used multiple models because there is inherent uncertainty in the choice of model that best predicts death rates in the absence of pandemic. These models were formulated to incorporate features of weekly death rates as follows:

- First, death rates may have a medium-to-long-term trend that affect mortality in 2020 compared to earlier years. We developed two sets of models, one with no trend and one with a linear trend term over weekly deaths.
- Second, death rates have a seasonal pattern, which varies by age group and sex^46-49^. We included weekly random intercepts for each week of the year. To account for the fact that seasonal patterns “repeat” (i.e., late December and early January are seasonally similar) we used a seasonal structure^50,51^ for the random intercepts. The seasonal structure allows the magnitude of the random intercepts to vary over time, and implicitly incorporates time-varying factors such as annual fluctuations in flu season.
- Third, death rates in each week may be related to rates in preceding week(s), due to short-term phenomena such as severity of the flu season. We formulated four sets of models to account for this relationship. The weekly random intercepts in these models had a first, second, fourth or eighth order autoregressive structure^50,51^ The higher-order autoregressive models allow death rates in any given week to be informed by those in a progressively larger number of preceding weeks. Further, trends not picked up by the linear or seasonal terms would be captured by these autoregressive terms.
- Fourth, beyond having a seasonal pattern, death rates depend on temperature, and specifically on whether temperature is higher or lower than its long-term norm during a particular time of year^52-57^. The effect of temperature on mortality varies throughout the year, and may be in opposite directions for different times of year. We used two sets of models, one without temperature and one with a weekly term for temperature anomaly, defined as deviation of weekly temperature from the local average weekly temperature over the entire analysis period. The coefficients of temperature anomalies were specified as a random effect with a random walk prior of order one, so that temperature effect is more similar in adjacent weeks. The random effect had a circular structure so that late December and early January are treated as adjacent.
- Death rates may be different around major holidays such as Christmas and New Year. We included effects (as fixed intercepts) for the week containing Christmas and New Year in all countries. For England and Wales and Scotland, we also included effects for the week containing and the week after other public holidays, because reported death rates in weeks that contain a holiday were different from other weeks. This term was tested but not included for other countries because the effect was negligible.
- We also tested, but did not include, terms for the weeks that coincided with a change to and from daylight saving time because the effect was negligible.

These choices led to an ensemble^45,58^ of 16 short-term Bayesian mortality projection models (2 trend options × 4 autoregressive options × 2 temperature options). We used the time-series of weekly reported deaths from the beginning of time series of data through mid-February 2020 to estimate the parameters of each model, which was then used to predict death rates for subsequent weeks as estimates of the counterfactual death rates (i.e., if the pandemic had not occurred). For the projection period, we used recorded temperature so that our projections take into consideration actual temperature in 2020. This choice of training and prediction periods assumes that the number of deaths that are directly or indirectly related to the Covid-19 pandemic was negligible through mid-February 2020 in these countries, but it allows for impacts to have appeared in subsequent weeks.

We used weakly-informative log gamma priors on log precision with both shape and rate equal to 0.001. We tested the sensitivity of the results to the choice of prior through the use of penalized complexity priors and found that the results were similar. All models were fitted using integrated nested Laplace approximation (INLA)^59^, implemented in the R-INLA software (version 20.03). We took 1,000 draws from the posterior distribution of age-specific deaths under each of the 16 models, and pooled the 16,000 draws to obtain the posterior distribution of age-specific deaths if the Covid-19 pandemic had not taken place. This approach incorporates both the uncertainty of estimates from each model and the uncertainty in the choice of model. The reported credible intervals represent the 2.5^th^ and 97.5^th^ percentiles of the posterior distribution of the draws from the entire ensemble. We also report the posterior probability that an estimated increase in deaths corresponds to a true increase (or decrease). Posterior probability would be 50% in a country and/or week in which an increase is statistically indistinguishable from a decrease, and larger posterior probability indicates more certainty in an increase.

We did all analyses separately by sex and age group (0-64 years, 65+ years) because death rates, and how they are impacted by the pandemic, vary by age group and sex. To obtain estimates across age groups and both sexes, we summed draws from age-sex-specific estimates.

### Validation of no-pandemic baseline weekly deaths

We tested how well our model ensemble estimates the number of deaths expected had the pandemic not occurred by withholding data for 15 weeks starting from mid-February (i.e., the same projection period as done for 2020) for an earlier year and using the preceding time-series of data to train the models. In other words, we created a situation akin to 2020 for an earlier year. We then projected death rates for the weeks with withheld data, and evaluated how well the model ensemble projections reproduced the known-but-withheld death rates. We repeated this for three different years: 2017 (i.e., train model using data from January 2010 to mid-February 2017 and test for the subsequent 15 weeks), 2018 (i.e., train model using data from January 2010 to mid-February 2018 and test for the subsequent 15 weeks), and 2019 (i.e., train model using data from January 2010 to mid-February 2019 and test for the subsequent 15 weeks). We performed these tests for both sexes and age groups used in the analysis. We report the projection error (which measures systematic bias) and absolute forecast error (which measures any deviation from the data). Additionally, we report coverage of the projection uncertainty; if projected death rates and their uncertainties are well estimated, the estimated 95% credible intervals should cover 95% of the withheld data.

The results of model validation (Extended Data Table 3) show that the estimates of how many deaths would be expected had the pandemic not occurred from the Bayesian model ensemble were unbiased, with mean projection errors of 1% (between -3% and 5% in different age groups, sexes and years). The mean absolute error was between 4% and 9% in different age groups, sexes and years. 95% coverage, which measures how well the posterior distributions of projected deaths coincide with withheld data was >90% for all age groups, sexes and years, which shows that the posterior distribution is well estimated.

### Strengths and limitations

The main strength of our work is the development and application of a method to systematically and consistently use time-series data from 2010 to early 2020 to estimate how many deaths would be expected in the absence of pandemic. The models incorporated important features of mortality, including seasonality of death rates, how mortality in one week may depend on previous week(s) and the seasonally-variable role of temperature. This methodology not only allows more robust estimation of the total impacts of the pandemic, but also enables comparisons of excess deaths across countries on a real-time basis. The use of a modelling framework, as we have done, allowed us to make estimates by age group and sex, which, because of smaller numbers of deaths, may not be possible (or at least stable) otherwise. By modelling death rates, rather than simply the number of deaths as is done in most other analyses, we account for changes in population size and age structure. We used an ensemble of models which typically leads to more robust projections and better accounts for both the uncertainty associated with each individual model and that of model choice^58^.

The main limitation of our work is that we did not have data on underlying cause of death beyond the distinction between Covid-19 and non-Covid deaths. Having a breakdown of deaths by underlying cause will help develop cause-specific models and understand which causes have exceeded or fallen below the levels expected. We also could not access age-specific and/or sex-specific data for a number of other countries. Nor did we have data on total mortality by socio-demographic status to understand inequalities in the impacts of the pandemic beyond deaths assigned to Covid-19 as the underlying cause of death. Releasing these data will allow more granular analysis of the impacts of the pandemic, which can in turn inform resource allocation and a more targeted approach to mitigating both the direct and indirect effects of Covid-19, now and for future waves of the pandemic.

## Data Availability

Estimates of weekly excess deaths by country will be available from http://globalenvhealth.org/code-data-download/ upon publication of the paper. Input data on deaths, population and temperature will also be available from http://globalenvhealth.org/code-data-download/.

## Code availability

The computer code for the Bayesian model ensemble used in this work will be available at http://globalenvhealth.org/code-data-download/ upon publication of the paper.

## Author contributions

ME, VK and JEB designed the study. VK and JEB developed and tested statistical methods with input from TR, RMP and ME. VK, RMP, TR and JEB accessed, harmonised and analysed data. VK wrote computer code, conducted analysis and prepared results. ME, VK, JEB and JP-S wrote the first draft of the paper and other authors contributed to the paper.

## Author information

ME reports a charitable grant from the AstraZeneca Young Health Programme, and personal fees from Prudential, outside the submitted work. JP-S is vice-chair of the Royal Society for Public Health and reports personal fees from Novo Nordisk A/S and Lane, Clark & Peacock LLP, outside of the submitted work.

## Acknowledgements

We thank Laura Clearly and Rod Jackson for information about data from New Zealand and Giulia Mangiameli for help with background materials and references.

---

**Extended Data Table 1.** Number of excess deaths from any cause and deaths assigned to Covid-19 due from mid-February to end of May 2020, by country.

|  | Number of excess deaths (95% credible interval) |  |  | Number of deaths assigned to Covid-19 as underlying cause<sup>1</sup> |
| --- | --- | --- | --- | --- |
|  | Women | Men | Both sexes |  |
| Austria | 670 (-10-1,300) | 310 (-640-1,200) | 970 (-230-2,100) | 668 |
| Belgium | 3,900 (2,700-4,900) | 4,600 (3,200-6,000) | 8,500 (6,600-10,300) | 9,483 |
| Bulgaria | -560 (-1,500-300) | -440 (-1,780-650) | -1,010 (-2,770-460) | 140 |
| Denmark | 320 (-220-820) | 300 (-300-850) | 620 (-210-1,400) | 571 |
| England & Wales | 30,000 (25,500-34,400) | 27,200 (20,500-33,200) | 57,200 (48,800-64,900) | 47,104 |
| Finland | 240 (-320-650) | 280 (-300-800) | 520 (-330-1,200) | 316 |
| France | 11,000 (5,800-15,700) | 11,600 (5,100-18,200) | 22,600 (14,000-30,900) | 28,771 |
| Hungary | -550 (-1,930-580) | -300 (-2,190-1,300) | -880 (-3,240-1,200) | 524 |
| Italy | 23,300 (17,400-28,900) | 22,900 (14,700-31,000) | 46,200 (35,600-56,500) | 33,340 |
| Netherlands | 5,200 (3,700-6,400) | 3,500 (1,700-5,100) | 8,600 (6,200-10,800) | 5,951 |
| New Zealand | -280 (-550--20) | -160 (-530-190) | -440 (-900-0) | 22 |
| Norway | 160 (-190-460) | 100 (-350-490) | 250 (-330-750) | 236 |
| Portugal | 1,300 (190-2,400) | 1,500 (90-2,900) | 2,900 (1,000-4,700) | 1,396 |
| Scotland | 2,300 (1,700-2,800) | 2,300 (1,600-3,000) | 4,600 (3,700-5,500) | 3,914 |
| Spain | 22,500 (18,900-25,900) | 23,200 (18,500-27,600) | 45,700 (39,800-51,300) | 27,127 |
| Sweden | 3,000 (2,300-3,600) | 2,500 (1,600-3,400) | 5,500 (4,400-6,500) | 4,395 |
| Switzerland | 920 (290-1,500) | 400 (-420-1,200) | 1,300 (260-2,300) | 1,656 |

<sup>1</sup> Data are from Office for National Statistics for England and Wales, and the European Centre for Disease Prevention and Control (ECDC) for other countries (https://www.ecdc.europa.eu/en/publications-data/download-todays-data-geographic-distribution-covid-19-cases-worldwide; accessed on 13 July 2020). ECDC data were only available for both sexes combined.
<sup>2</sup> Data on all-cause mortality, which is used to estimate excess deaths, for Italy are from municipalities with up-to-date death registration, that cover 95.0% of the population and 98.0% of COVID deaths. Data for deaths assigned to Covid-19 are for the entire country.

**Extended Data Table 2.**
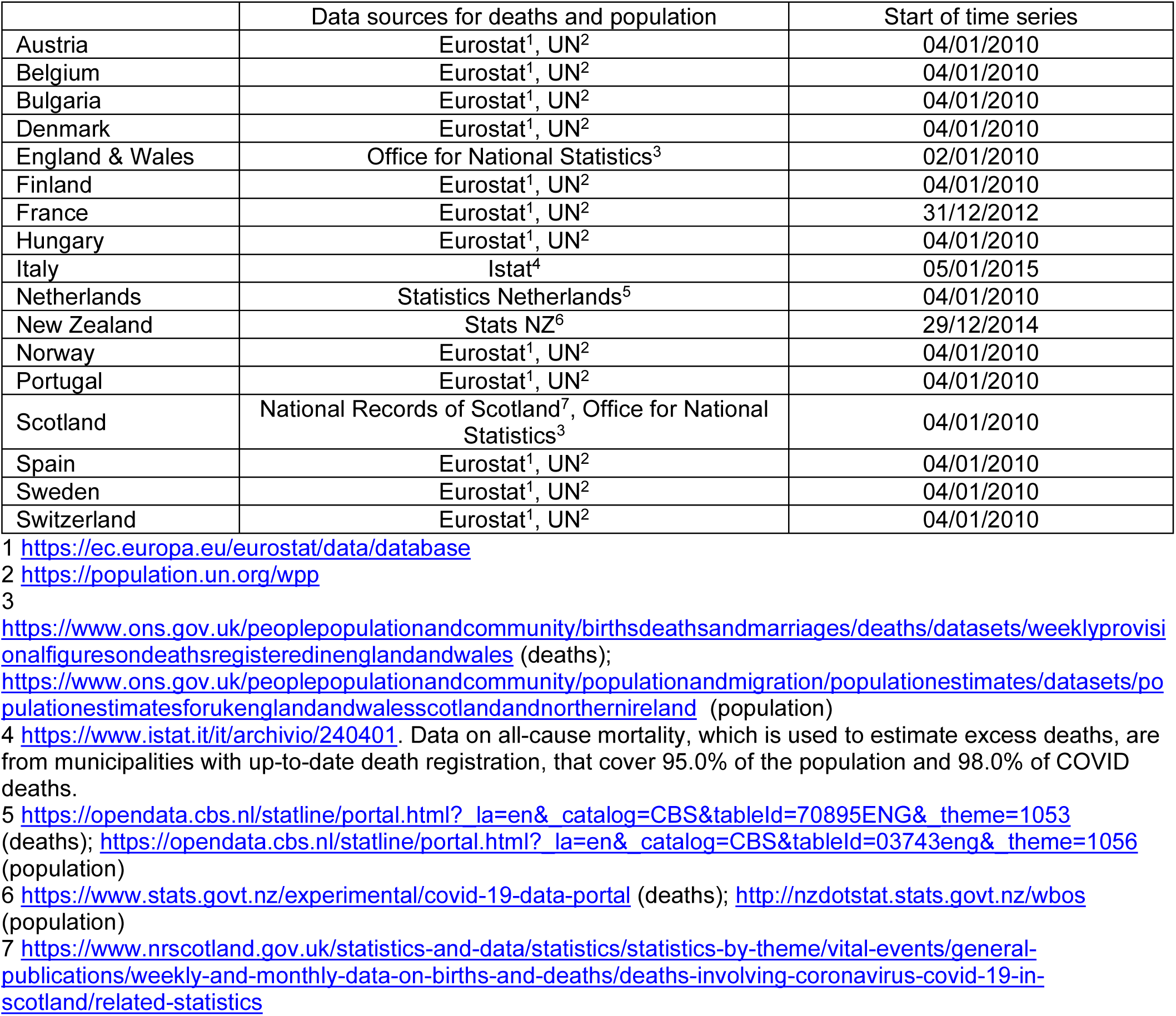
Sources of data on death and population used in the paper.

|  | Data sources for deaths and population | Start of time series |
| --- | --- | --- |
| Austria | Eurostat<sup>1</sup>, UN<sup>2</sup> | 04/01/2010 |
| Belgium | Eurostat<sup>1</sup>, UN<sup>2</sup> | 04/01/2010 |
| Bulgaria | Eurostat<sup>1</sup>, UN<sup>2</sup> | 04/01/2010 |
| Denmark | Eurostat<sup>1</sup>, UN<sup>2</sup> | 04/01/2010 |
| England & Wales | Office for National Statistics<sup>3</sup> | 02/01/2010 |
| Finland | Eurostat<sup>1</sup>, UN<sup>2</sup> | 04/01/2010 |
| France | Eurostat<sup>1</sup>, UN<sup>2</sup> | 31/12/2012 |
| Hungary | Eurostat<sup>1</sup>, UN<sup>2</sup> | 04/01/2010 |
| Italy | Istat<sup>4</sup> | 05/01/2015 |
| Netherlands | Statistics Netherlands<sup>5</sup> | 04/01/2010 |
| New Zealand | Stats NZ<sup>6</sup> | 29/12/2014 |
| Norway | Eurostat<sup>1</sup>, UN<sup>2</sup> | 04/01/2010 |
| Portugal | Eurostat<sup>1</sup>, UN<sup>2</sup> | 04/01/2010 |
| Scotland | National Records of Scotland<sup>7</sup>, Office for National Statistics<sup>3</sup> | 04/01/2010 |
| Spain | Eurostat<sup>1</sup>, UN<sup>2</sup> | 04/01/2010 |
| Sweden | Eurostat<sup>1</sup>, UN<sup>2</sup> | 04/01/2010 |
| Switzerland | Eurostat<sup>1</sup>, UN<sup>2</sup> | 04/01/2010 |

1 https://ec.europa.eu/eurostat/data/database
2 https://population.un.org/wpp
https://www.ons.gov.uk/peoplepopulationandcommunity/birthsdeathsandmarriages/deaths/datasets/weeklyprovisionalfiguresondeathsregisteredinenglandandwales (deaths);
https://www.ons.gov.uk/peoplepopulationandcommunity/populationandmigration/populationestimates/datasets/populationestimatesforukenglandandwalesscotlandandnorthernireland (population)
4 https://www.istat.it/it/archivio/240401. Data on all-cause mortality, which is used to estimate excess deaths, are from municipalities with up-to-date death registration, that cover 95.0% of the population and 98.0% of COVID deaths.
5 https://opendata.cbs.nl/statline/portal.html?_la=en&_catalog=CBS&tableId=70895ENG&_theme=1053 (deaths); https://opendata.cbs.nl/statline/portal.html?_la=en&_catalog=CBS&tableId=03743eng&_theme=1056 (population)
6 https://www.stats.govt.nz/experimental/covid-19-data-portal (deaths); http://nzdotstat.stats.govt.nz/wbos (population)
7 https://www.nrscotland.gov.uk/statistics-and-data/statistics/statistics-by-theme/vital-events/general-publications/weekly-and-monthly-data-on-births-and-deaths/deaths-involving-coronavirus-covid-19-in-scotland/related-statistics

**Extended Data Table 3.** Results of the external predictive validity (out-of-sample validation) of the estimated no-pandemic baseline weekly deaths from the Bayesian model ensemble. Each number represents the total error over the validation period, averaged across countries.

| Validation year | Age group (years) | Projection error (relative projection error) |  | Absolute projection error (relative absolute projection error) |  | Percent covered by 95% credible interval |  |
| --- | --- | --- | --- | --- | --- | --- | --- |
|  |  | Men | Women | Men | Women | Men | Women |
| 2017 | 0-64 | 101 (2%) | 10 (1%) | 229 (7%) | 165 (9%) | 96% | 98% |
|  | 65+ | 703 (4%) | 945 (5%) | 892 (6%) | 1112 (6%) | 95% | 94% |
| 2018 | 0-64 | -90 (-3%) | -36 (-2%) | 230 (8%) | 171 (9%) | 96% | 97% |
|  | 65+ | -201 (-2%) | -240 (-1%) | 1045 (7%) | 1392 (8%) | 87% | 89% |
| 2019 | 0-64 | 86 (3%) | 45 (2%) | 236 (8%) | 166 (9%) | 96% | 95% |
|  | 65+ | 369 (2%) | 473 (2%) | 670 (4%) | 857 (4%) | 97% | 100% |
| All three years | 0-64 | 33 (1%) | 6 (0%) | 232 (8%) | 168 (9%) | 96% | 97% |
|  | 65+ | 290 (1%) | 393 (2%) | 869 (6%) | 1120 (6%) | 93% | 94% |

\* France, Italy and New Zealand were not used for validation analysis because they had shorter time series. Hence leaving out the last three years of data would leave a time series that was too short for estimating model parameters.

**Extended Data Figure 1.**
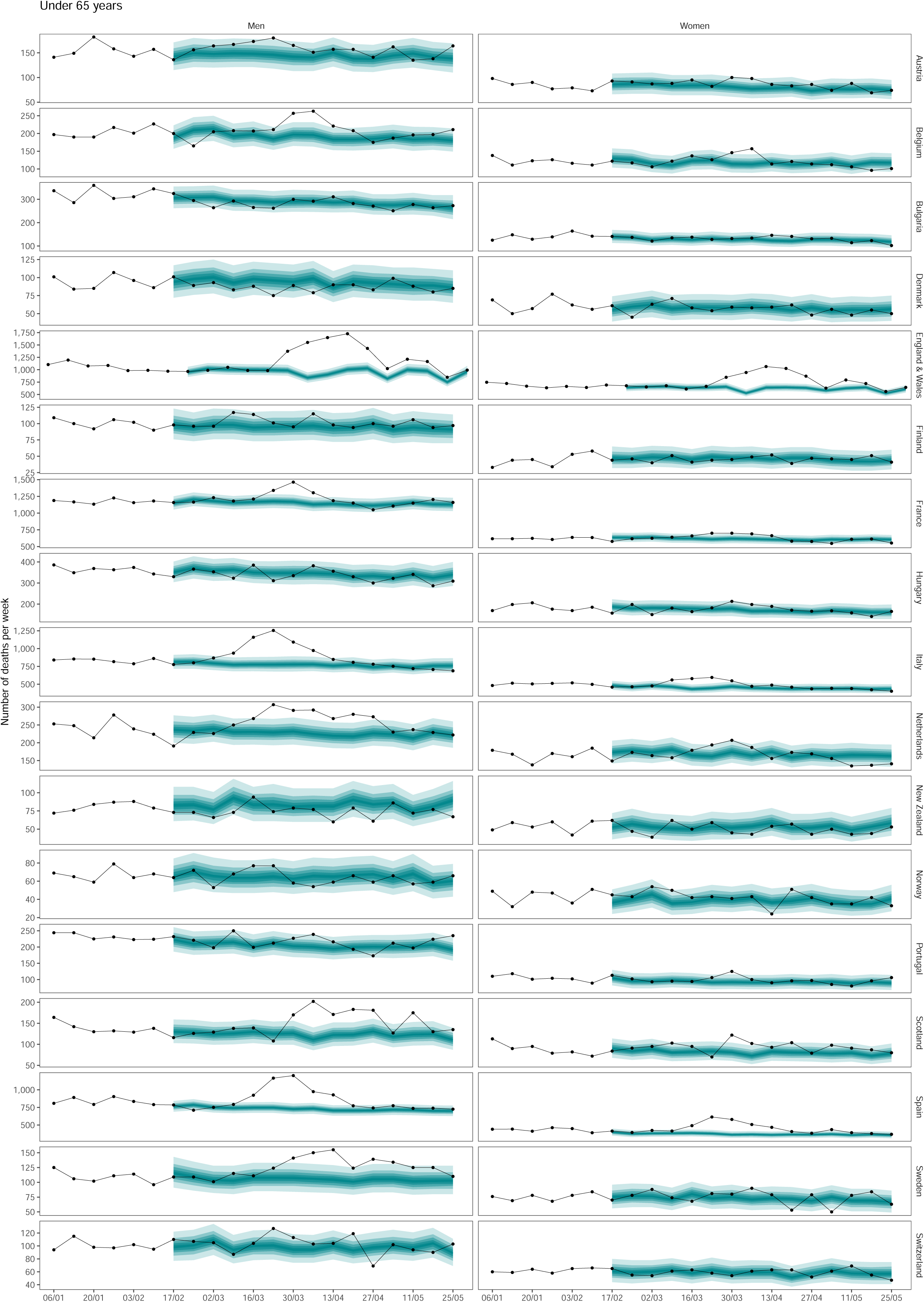
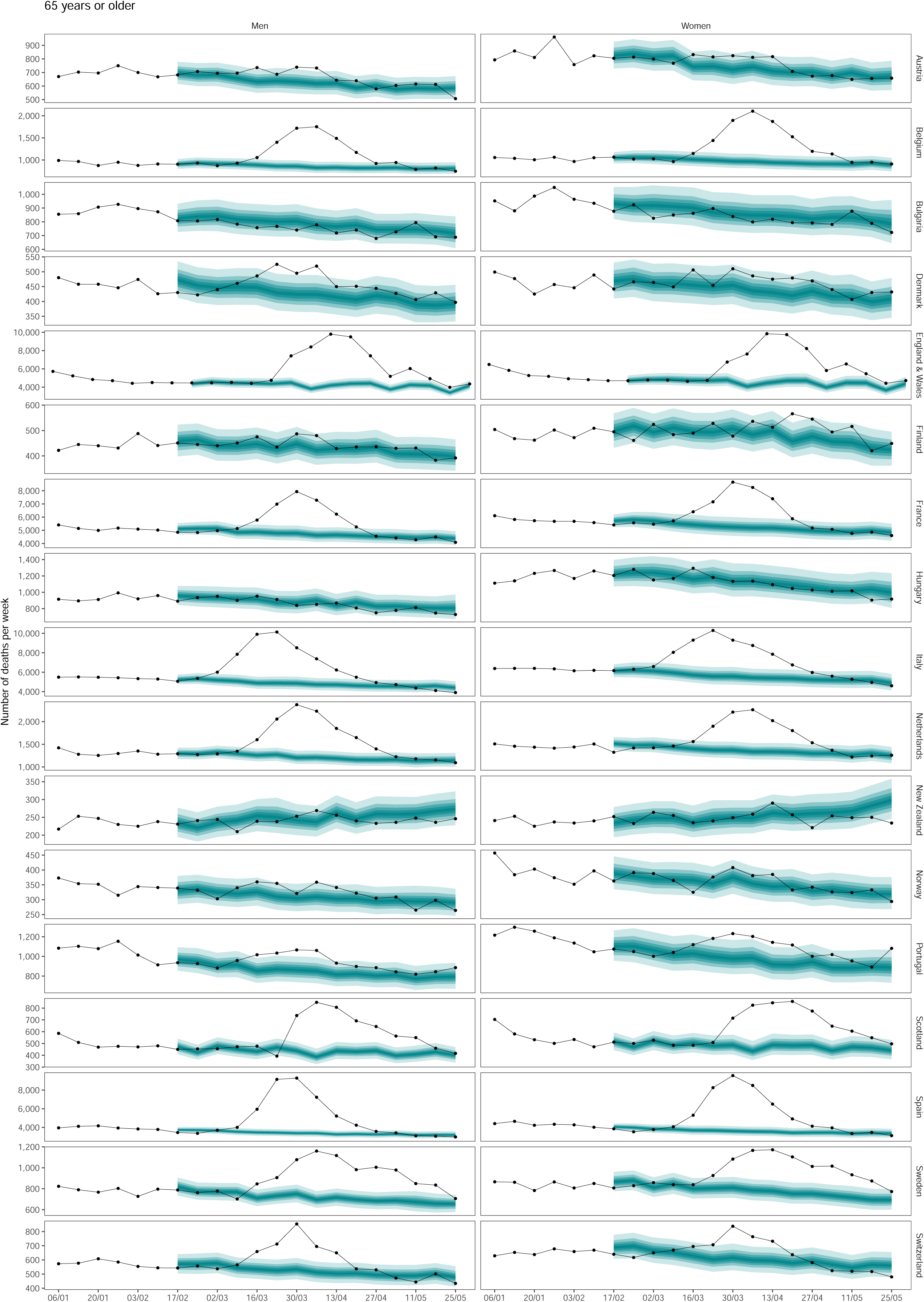
Weekly number of deaths from January 2020 through May 2020, by age group. The points show reported deaths. The grey-shaded areas show the predictions of how many deaths would have been expected from mid-February had Covid-19 pandemic not taken place. The turquoise shading shows the credible intervals around the median prediction, from 5% (dark) to 95% (light) in 10% increments.

**Extended Data Figure 2.**
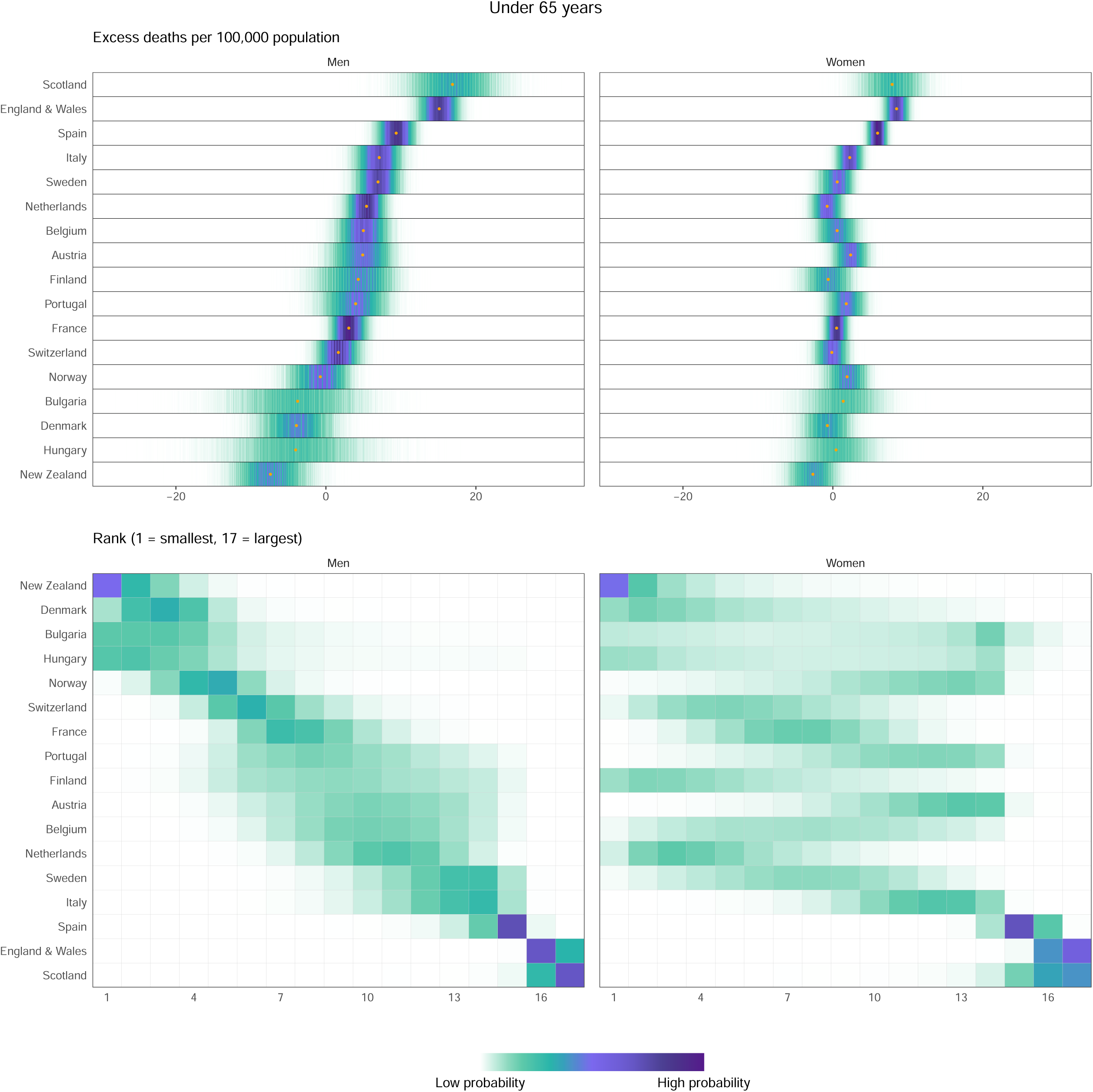
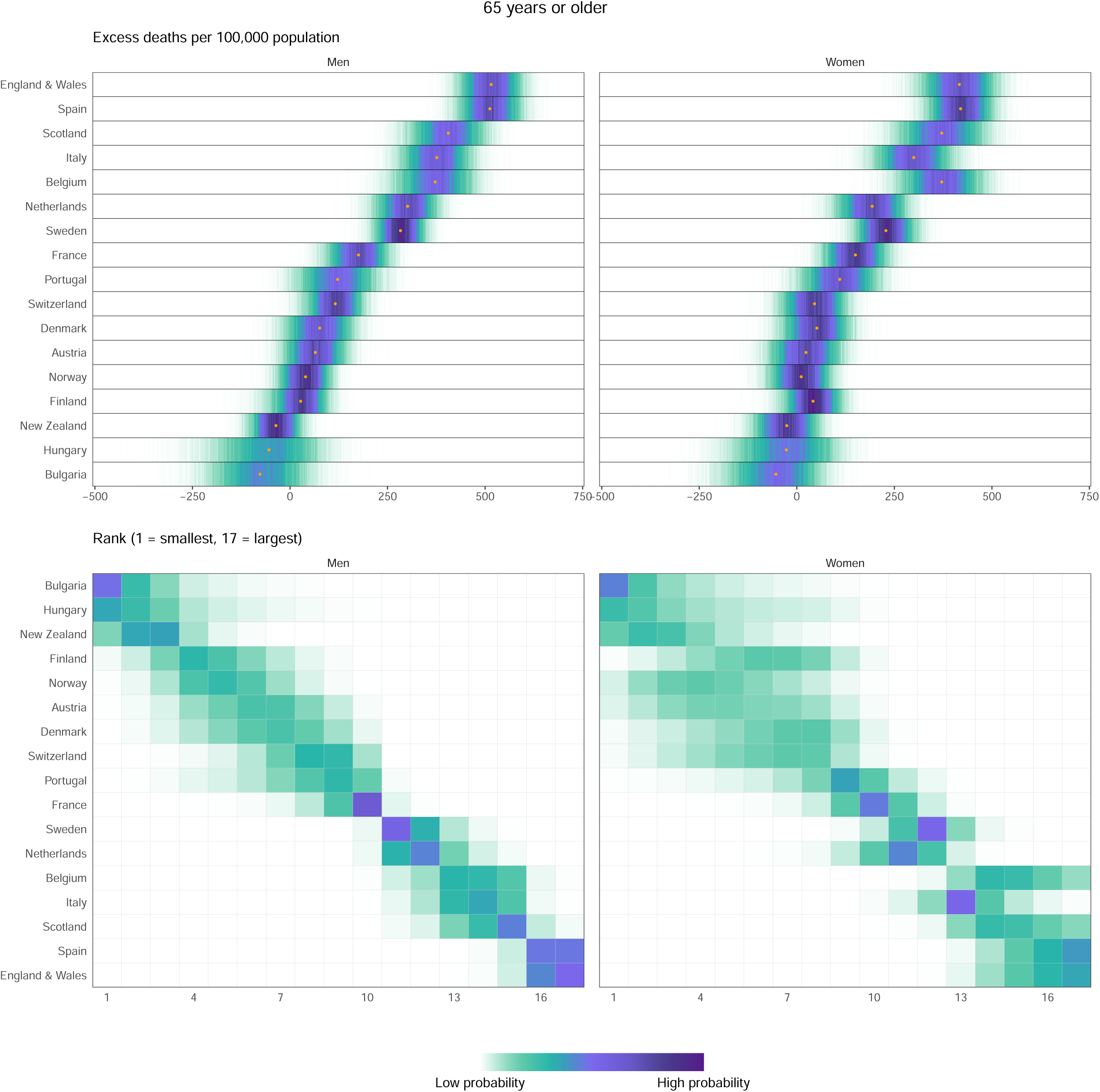
(A) Posterior distribution of excess deaths from any cause per 100,000 people from mid-February to end of May 2020, by age group. Gold dots show the posterior medians. (B) Probability distribution for the country’s rank, by age group. Countries are ordered vertically by median increase from smallest (at the bottom) to the largest (at the top).

**Extended Data Figure 3.**
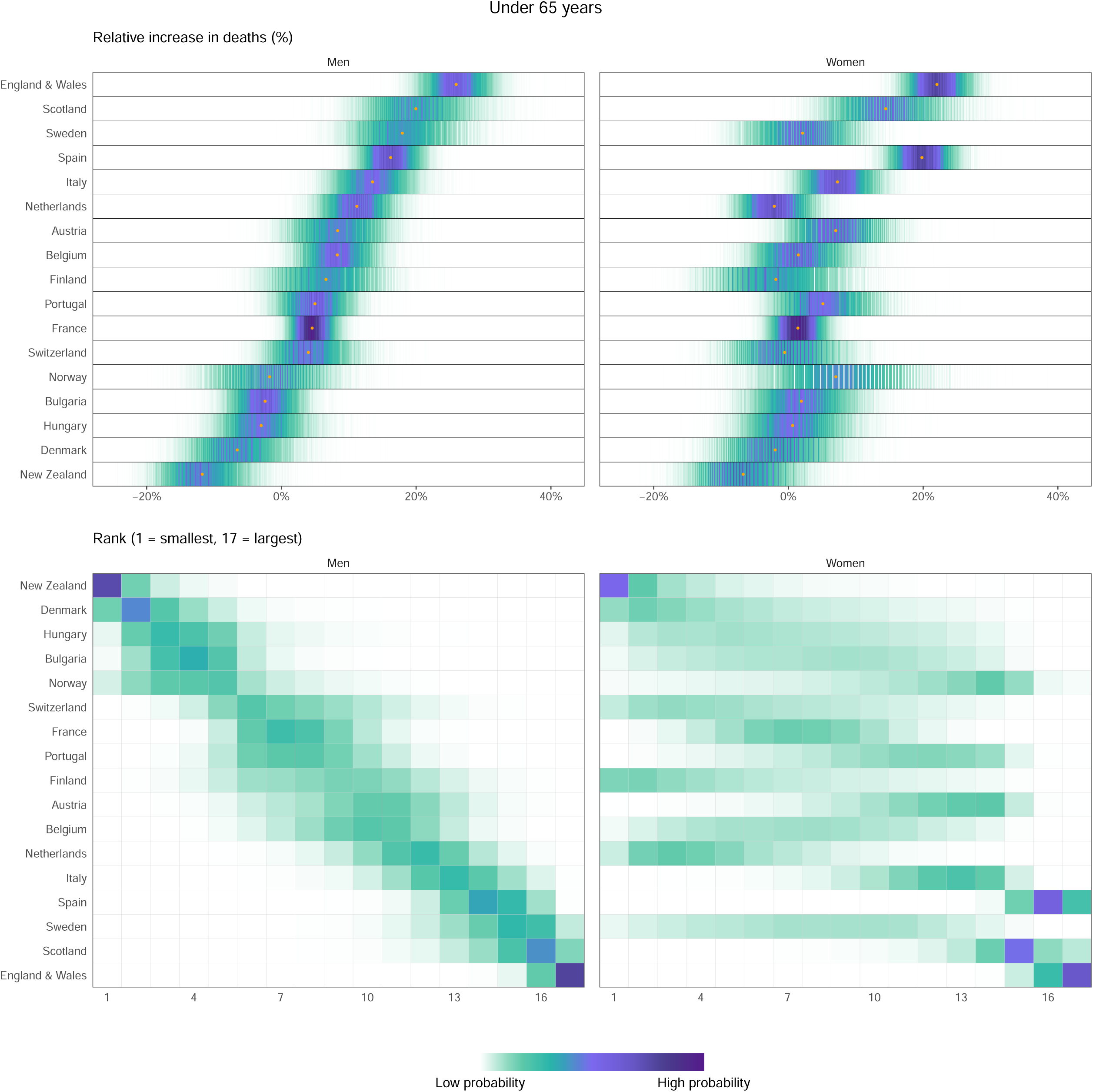
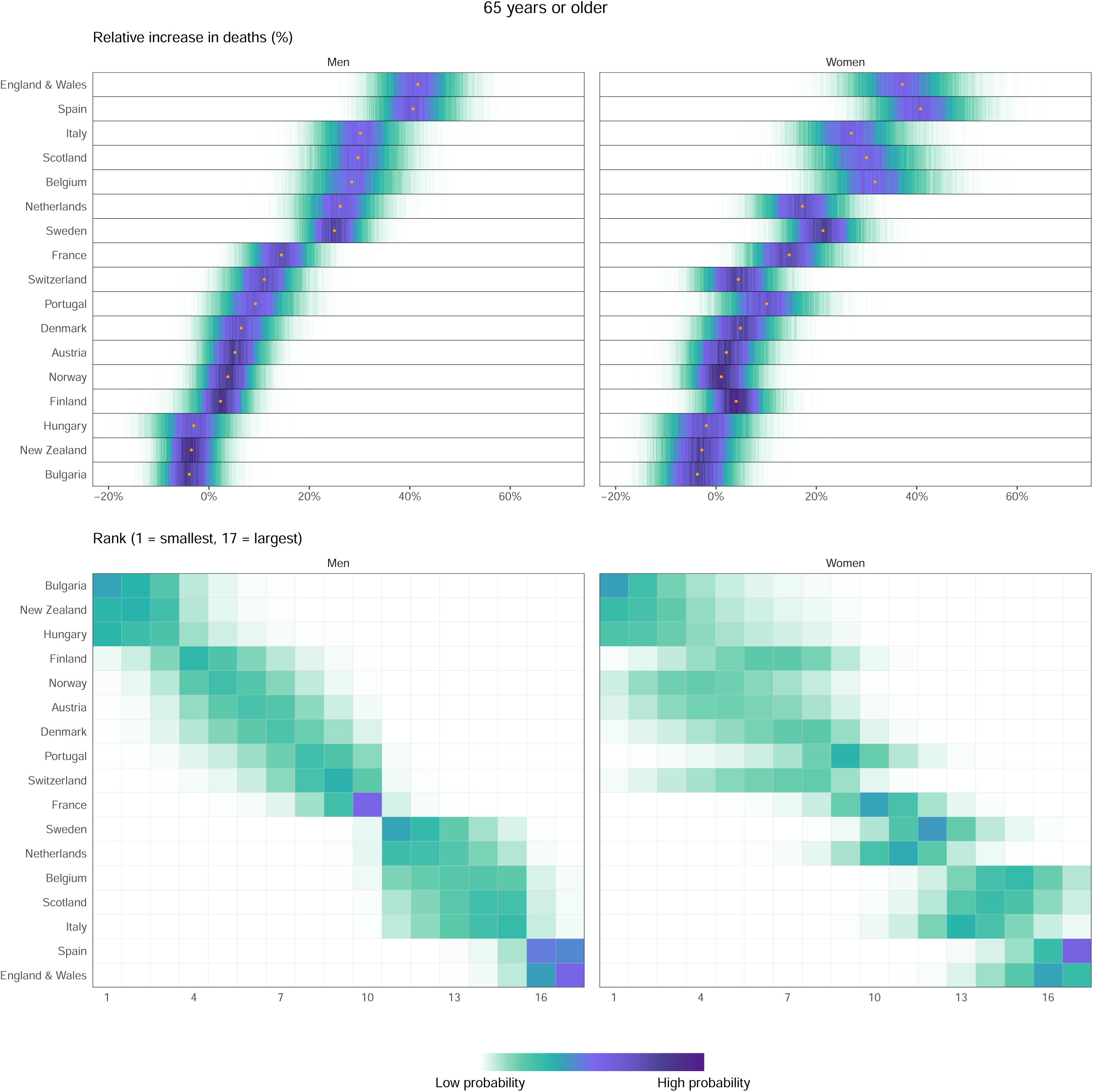
(A) Posterior distribution of percent increase in deaths from any cause from mid-February to end of May 2020, by age group. Gold dots show the posterior medians. (B) Probability distribution for the country’s rank, by age group. Countries are ordered vertically by median increase from smallest (at the bottom) to the largest (at the top).

**Extended Data Figure 4.**
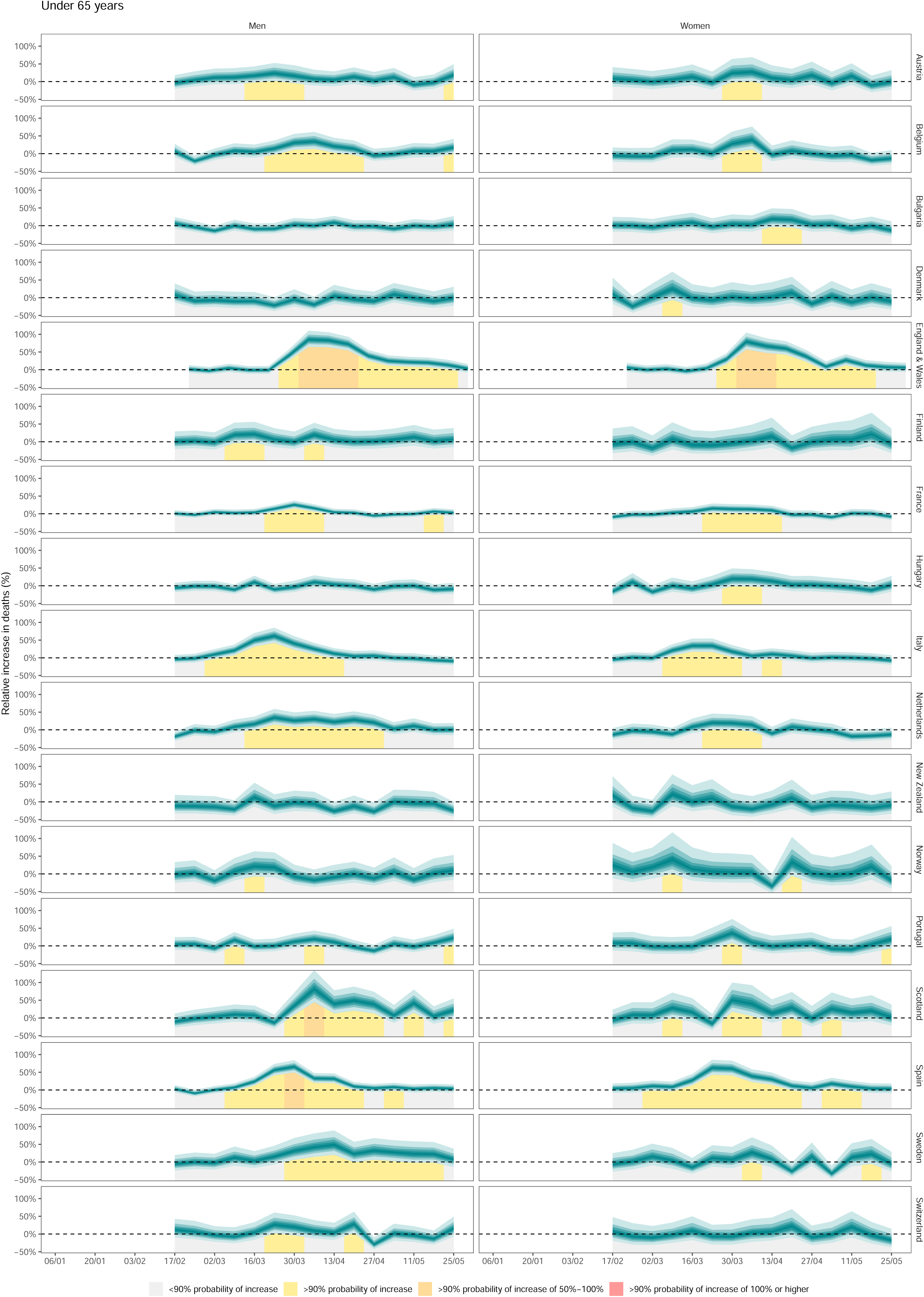
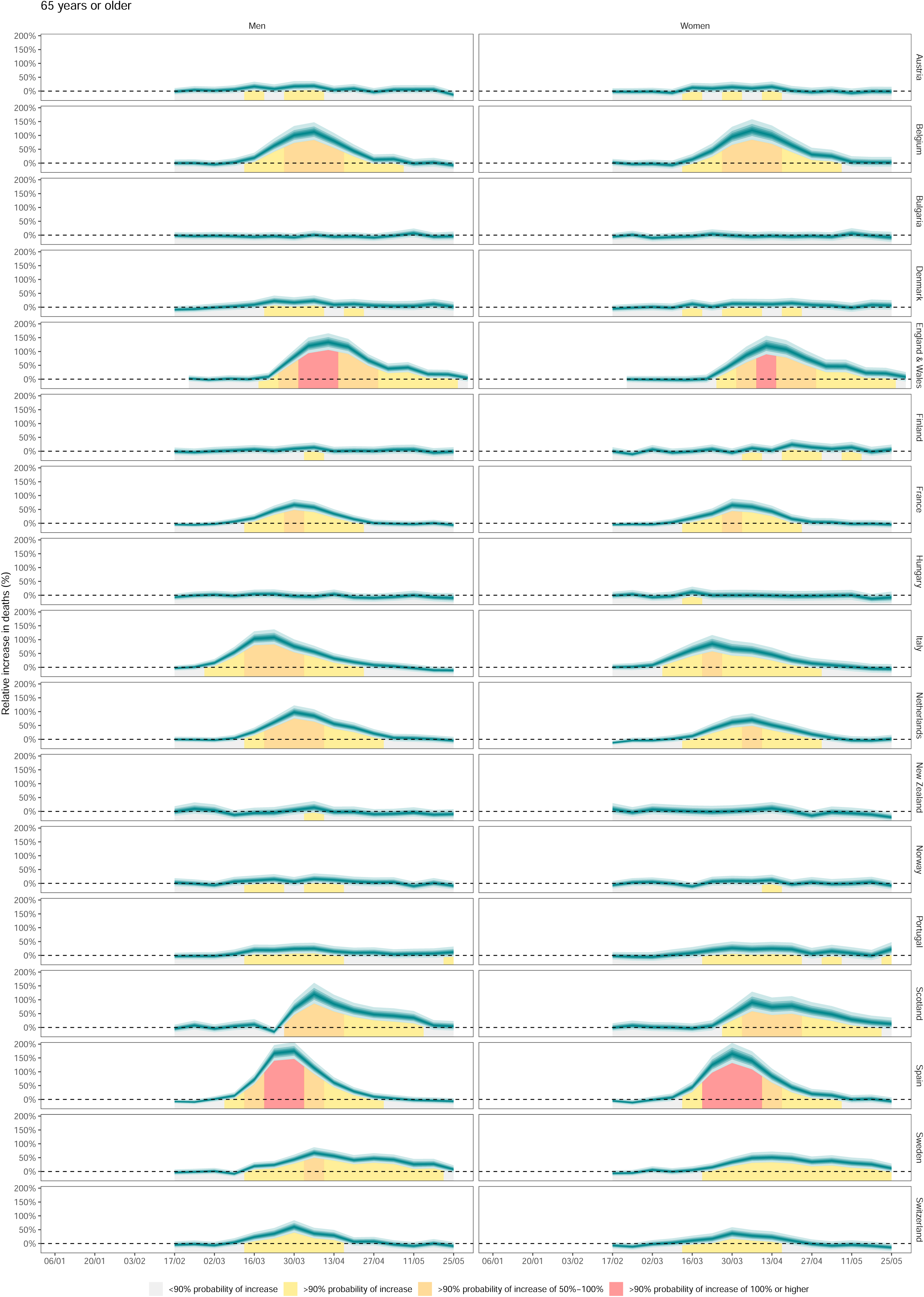
Weekly percent increase in mortality as a result of the Covid-19 pandemic by country, by age group. The turquoise shading shows the credible intervals around the median prediction, from 5% (dark) to 95% (light) in 10% increments. The background shading indicates the magnitude of the weekly increase that was detectable with a posterior probability of at least 90%.

## Notes

### Funding Statement

No external funding was received for this work.

